# A secure and rapid query-software for COVID-19 test results that can easily be integrated into the clinical workflow to avoid communication overload

**DOI:** 10.1101/2020.04.07.20056887

**Authors:** Gunnar Völkel, Axel Fürstberger, Julian D. Schwab, Silke D. Kühlwein, Thomas Gscheidmeier, Johann M. Kraus, Alexander Groß, Florian Kohlmayer, Peter Kuhn, Klaus A. Kuhn, Oliver Kohlbacher, Thomas Seufferlein, Hans A. Kestler

## Abstract

Overcoming the COVID-19 crisis requires new ideas and strategies. Rapid testing of a large number of subjects is essential to monitor, and delay, the spread of SARS-CoV-2 to mitigate the consequences of the pandemic. People not knowing that they are infected may not stay in quarantine and, thus, are a risk for infecting others. Unfortunately, the massive number of COVID-19 tests performed is challenging for both laboratories and the units that take the throat swab and have to communicate test results. Here, we present a secure tracking system (CTest) to report COVID-19 test results online as soon as they become available. The system can be integrated into the clinical workflow with very modest effort and avoids excessive load to telephone hotlines. With this open-source and browser-based online tracking system, we aim to minimize the time required to inform the tested person but also the test units, e.g. hospitals or the public healthcare system. Instead of personal calls, CTest updates the status of the test automatically when the test results are available. Test reports are published on a secured web-page enabling regular status checks also by patients not using smartphones with dedicated mobile apps which has some importance as smartphone usage diminishes with age.

The source code, as well as further information to integrate CTest into the IT environment of other clinics or test-centres, are freely available from https://github.com/sysbio-bioinf/CTest under the Eclipse Public License v2.0 (EPL2).

## Introduction

After the first outbreak of SARS-CoV-2 in Wuhan (Hubei province, China) in December 2019, the has been spreading rapidly around the world [1]. Thus, the management of its induced crisis has become a ubiquitous topic [2]. Each day, the number of new infections increases dramatically worldwide reaching globally 1,276,302 confirmed cases on the April 4th 2020 (9:28 AM) [3] with a median age of 51 years [1]. The most common test for COVID-19 infections is to take a throat swab and test by real-time polymerase chain reaction (RT-PCR) [1,4].

Infected persons can spread the virus before the first symptoms appear [5,6]. Timely communication of results is thus essential to take appropriate action, but challenging due to the throughput and the often demanding circumstances tests are carried out. Current practice in Germany is to call the respective clinic or laboratory for information about test results. This process ties up considerable resources and does not scale well for large numbers of tests. Communication channels become overloaded due to repeated calls of the testees and waiting times as well as anxiety of the testee’s increase.

To accelerate the COVID-19 test procedure, we implemented an online query system, called CTest. It empowers testees since they can check their test result status independently through a convenient, secure online system (see Fig 1) [7]. The approach avoids unnecessary and repeated phone calls, manual transcription errors, and consequently reduces the burden on the clinical staff.

**Fig 1.**
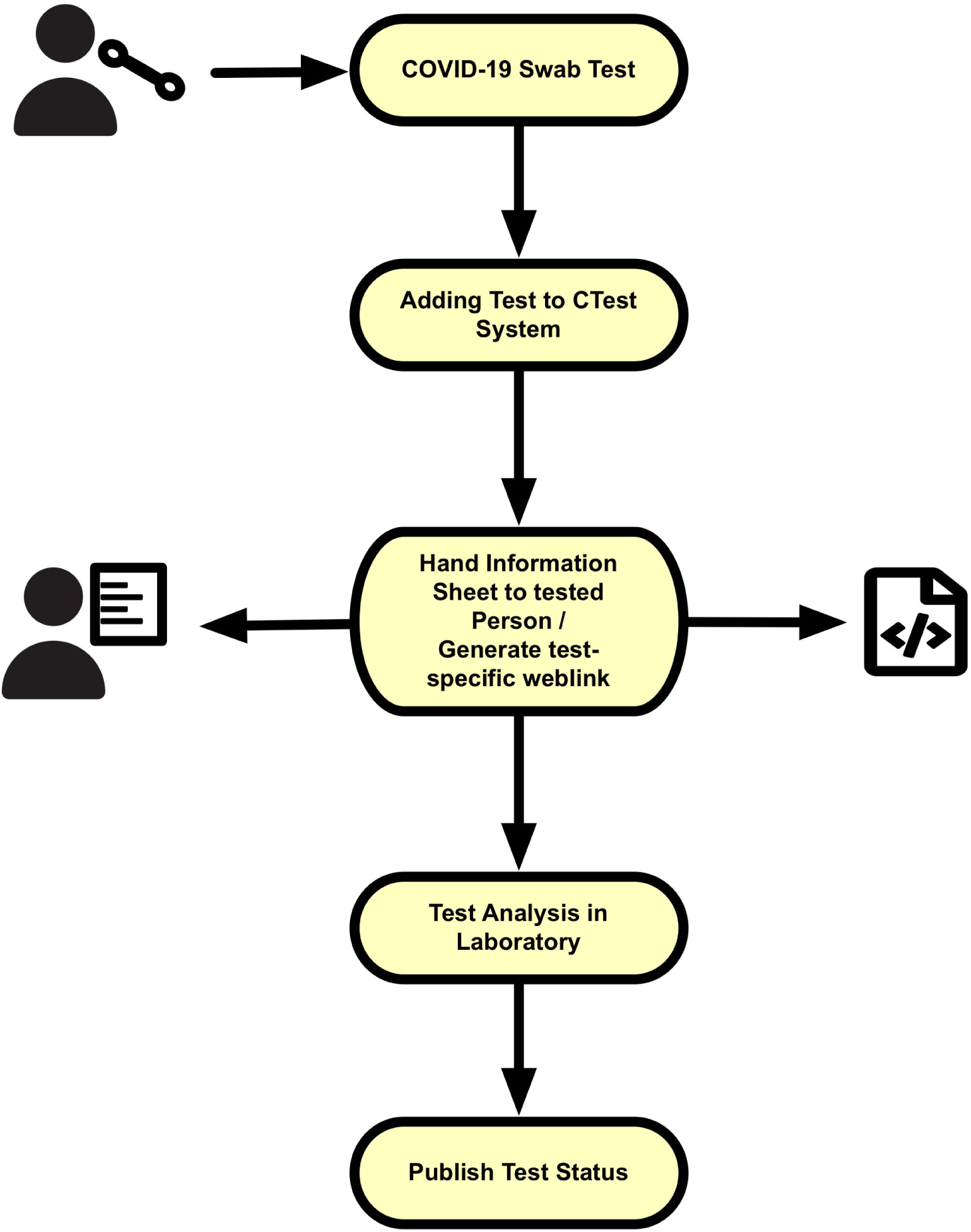
Workflow of the COVID-19 test process using CTest. First, COVID-19 swab tests are extracted. The test is then added to the CTest system via an order number. Based on this order number CTest generates a cryptographically secure tracking-ID. An information sheet with a test-specific weblink is generated by CTest and can be handed over to the person tested. After laboratory analysis is complete, the results are sent to CTest. CTest updates the test results that can be queried via the individual web link.

The CTest system extends the previously developed online tracking tool TraqBio [8]. This application was created to simplify and standardise communication between users and CORE facilities. Its clean design and the open-source license allow development (refactoring), deployment, establishment, and integration of CTest within a short period of time.

CTest runs on the Java Virtual Machine [8,9]. The web application can be deployed independently of operating system and platform.

## Methods

### Implementation and setup

We based the CTest application on the TraqBio software [8]. Backend functionality of the CTest server is implemented in the Lisp dialect Clojure. Clojure runs on the Java Virtual Machine [9] and is thus platform-independent. The backend comprises a database to store the scheduled COVID-19 tests and corresponding results. Clojure supports different databases and connectors. We choose SQLite (version 3.29.0) [10] to keep the setup independent of additional database servers. A local database file holds the data. Additionally, CTest provides functionality for user management and data backup.

Every new order number entered and supplied by the laboratory staff in the CTest system, generates a database entry with a unique identifier and unique tracking number. Here, this order number is used as the primary unambiguous identifier. CTest can be configured to append the current date to the order number to create a primary unambiguous identifier in setups where order numbers are only guaranteed to be unique within the same day. Using a secure random number generator, we generate a corresponding tracking number [11]. This random number generator generates six bytes which are then translated into a sequence of 12 characters between 0-9 and A-F. This sequence of characters is unique and not created in sequential order. Every tracking number generated is checked for uniqueness prior to use; there is no link between tracking and order number.

Web-frontend functionality is implemented using Javascript and HTML templates. We used freely available standard web frameworks like bootstrap (v3) [12, 13], jQuery [14] and extensions to these frameworks. Using these state-of-the-art frameworks, we aim to enable a straight-forward adaptation of the frontend for integration at other institutions. The frontend features a responsive graphical user interface for the management functions, the creation of tests and, accessibility for users to their test results. The test status can be queried via a unique weblink without requiring an account or login.

To secure web communication, creation, and queuing our setup consists of different security layers. Only Secure Sockets Layer (SSL) certified access to the websites is allowed via Hypertext Transfer Protocol Secure (HTTPS). Connection and transferred data are encrypted, and the CA-signed server certificates are used. A reverse proxy setup forwards the external hostname to a virtual machine within the hospital’s secured network infrastructure. On the virtual server, another reverse proxy is in place to allow running the Java application as a non-privileged user. As the standard http port 80 is privileged, it can only be used by a system user. So, running the application as a user with system-wide rights is a security risk. Therefore, proxy settings forward the privileged port to a high port (above 1024), that normal users can control. Hence, no system rights are necessary for managing the application on the operating system level. For the setup in the clinical environment, we also set up a firewall with specific ban rules (iptables and fail2ban service on a Linux operating system) to prevent brute force attacks on login or tracking numbers. Also, network IPs and subnets can be white-listed to allow access to management functions of the application. Thus, other computers and devices are blocked from accessing these functions after failed attempts. For the tracking interface, a brute force attack (like trying all combinations of possible tracking numbers) is shielded by blocking IPs after too many failed attempts with wrong or non-existing tracking numbers.

### Query performance test

For our performance test, we first performed 1024 simultaneous requests using a single machine and a single network connection. Next, we created a mixed dataset (interleaved_urls), including 758 available database entries (available_urls) and the same number of non-available tracking numbers (notavailiable_urls). Furthermore, we measured queries for always the same one or two URLs. That makes in total six datasets, each of which was accessed 1516 times. Queries were carried out once in sequence (ordered) and randomly (random). Also, the two scenarios caching function of the browser (1filePerRequest) and complete reloading (23or1filesPerRequest) were tested. Meaning that once unchanged files are not reloaded (caching), whereas in the other scenario, all required files and displayed images (e.g. flag-graphics or css) are reloaded. For the second scenario, one file is loaded if the tracking number is not available and 23 files if the tracking number is available. Stress tests were measured using Siege 4.0.4 [15].

## Results

CTest was built based on an existing, proven software stack (TraqBio [8]). It extends TraqBio to the functionality required for COVID-19 tests. We could successfully integrate it into the existing clinical testing workflows for SARS-CoV-2 infections in a major German university medical centre within a few days (Fig 2).

**Fig 2.**
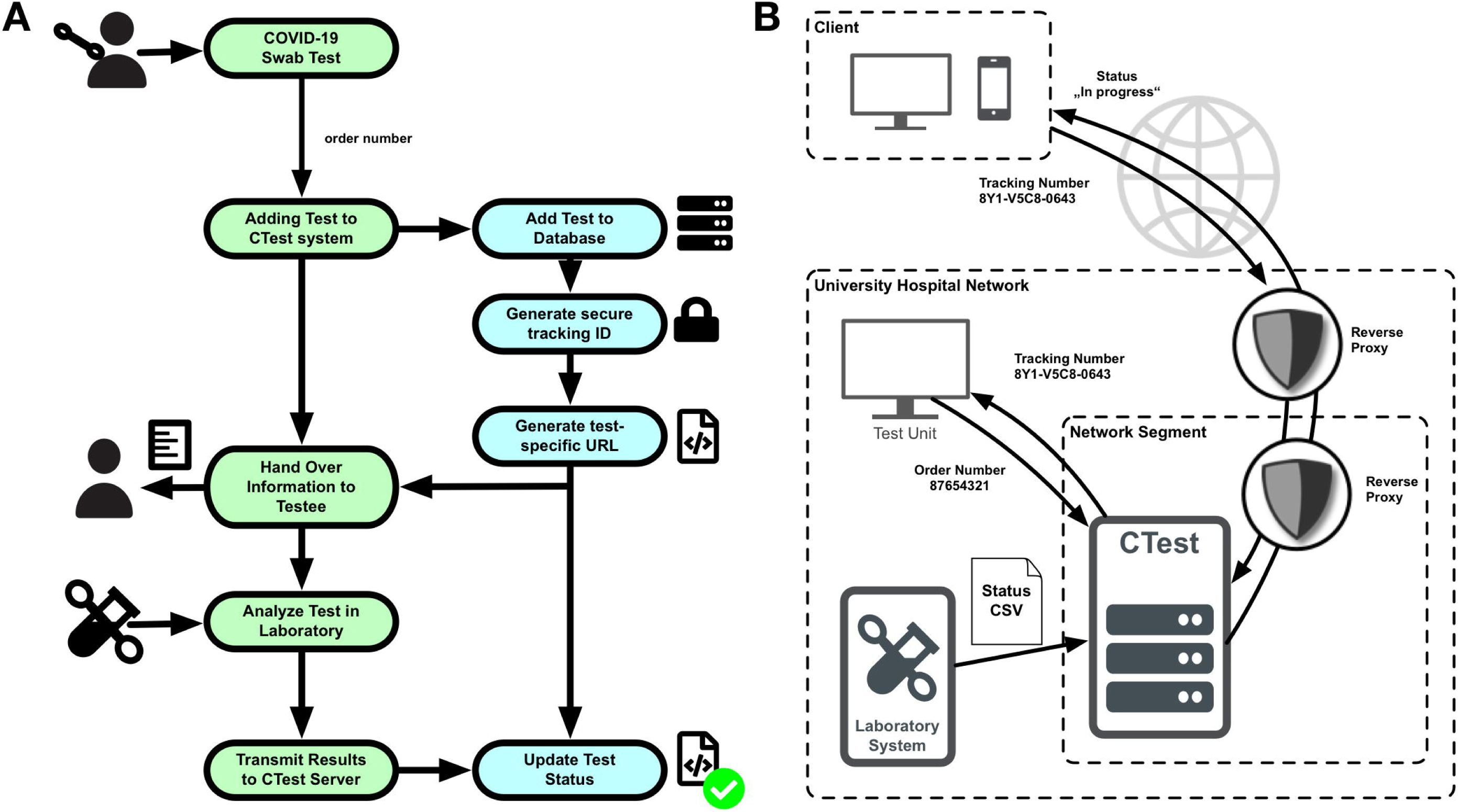
COVID-19 test procedure and set up. (A) The left panel shows the detailed process flow of the COVID-19 tests. CTest (blue) is tightly integrated into the test process (green). After the swab test for COVID-19, each test is included in the CTest database via a test-specific order number. An order number, e.g. from a laboratory information system, is used to generate a cryptographically secure tracking-ID. CTest then automatically creates a test-specific information sheet for each testee. This sheet contains a test-specific weblink and QR-code, based on the tracking-ID. The URL does not provide any information which allows for tracking back the test results to the corresponding testee. After the analysis in the laboratory, results are transmitted to the CTest server. CTest automatically updates the status on the test-specific weblink based on the corresponding test results. (B) In the right panel, a possible integration scenario of CTest into a clinical infrastructure is shown. CTest runs on a virtual machine within the secured network of a university hospital. The external hostname with web links to the testee’s statuses is forwarded via reverse proxy. A second reverse proxy forwards the port to the application to a non-privileged port. Thus, the application does not need to run with a system user. The test unit inside the hospital’s network communicates to the CTest server to create new test cases via an order number. The CTest sever returns back the tracking number and the corresponding weblink/QR-Code. The laboratory pushes test results as comma-separated-value format (CSV) files to the CTest server. The test-specific web page content is then updated according to the test result.

The workflow starts with taking a sample for testing. In a first step, an order number is generated by the testing lab and added as a barcode label to the sample. Analogous to other medical applications, order numbers within the laboratory information system of the clinical information system are unique but do not contain personal information about the patient [16]. This order number is transferred to the CTest system via scanning or typing the number into an input field of the dialogue window. We implemented format restrictions via regular expressions to the input field to minimize incorrect entries. Afterwards, CTest generates an unambiguous, non-consecutive tracking number. Therefore, a cryptographically strong pseudo-random number generator [11] creates six bytes that are transferred into a twelve-digit character code, including letters A-F and numbers from 0 to 9. Using this tracking number ensures that no personal information about any testee can be inferred.

After taking the sample, the tracking number is given to each testee on a printed sheet, including information on how to access the status of their COVID-19 test (Fig 3A). After the sample has been processed, the lab system sends update files as a comma-separated-value (CSV) file via secure encrypted file transfer protocol-connection (SFTP) to the CTest server. Results in the CSV file are then automatically parsed, backed up, and imported into the database of CTest which leads to a fully automated update of the status of each processed test.

**Fig 3.**
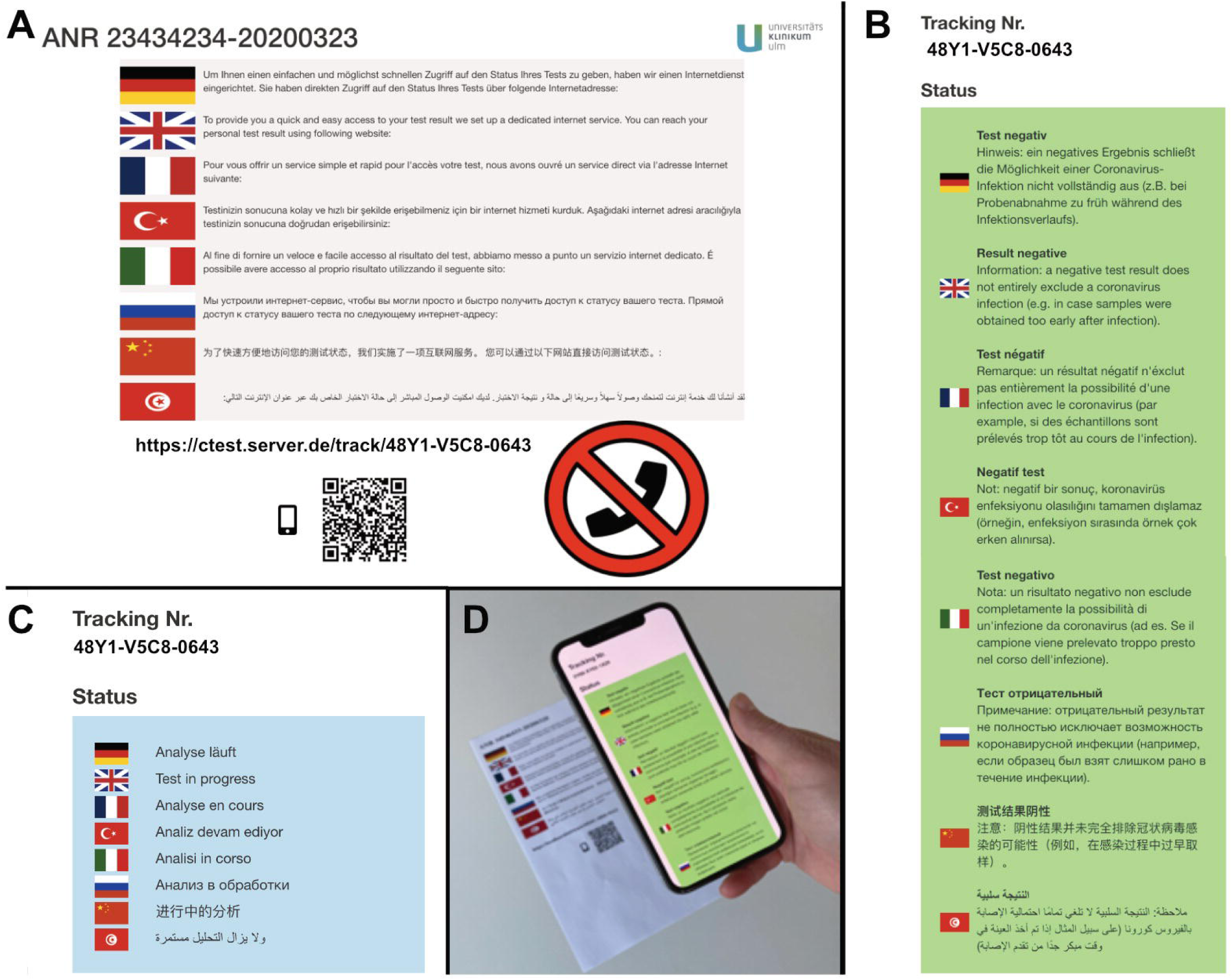
Information for testees. Information concerning the access to the test status and its results are given in eight languages. (A) The information sheet that is handed over to testees. It informs how to access the COVID-19 test results online. The test-specific URL links to a page which includes information about negative test result (B) or that the test is still in progress (C). Scanning the QR code with a smartphone forwards directly to the current test status or its result (D).

Currently, the CTest system distinguishes two potential outcomes: 1) the COVID-19 test is negative (Fig 3B) or 2) still in progress (Fig 3C). Positive results are mapped to “in progress”. Personal phone calls from the health department will inform people who are tested positive for SARS-CoV-2. Thereby they can be informed about health arrangements and how to avoid further spreading of the virus.

We provide two possibilities to query the status of the COVID-19 test. Either it is possible to scan a QR code on the received information letter with a smartphone to get redirected to a test status webpage (Fig 3D) or to enter this weblink into a web browser directly. The status of the individual test result is automatically displayed in a responsive form on the device (Fig 3D). Currently, we observe that both possibilities to request the test results are used. 30.6% of people prefer a query of the test results via a web browser while the rest of testees scan the provided QR-code.

To overcome language barriers, information concerning the procedure to obtain test results and the results were translated into eight languages, including German, English, French, Turkish, Italian, Russian, Chinese, and Arabic.

Due to the high number of tests being administered during the peak times of infection waves, CTest was designed to handle large numbers of queries in a short amount of time. Since the introduction of Ctest into the clinical routine, we detect around 12 queries per performed test (Fig 4A). Handling of all these requests by phone calls would lead to communication overload. Consequently, CTest can reduce the burden on clinical staff.

**Fig 4.**
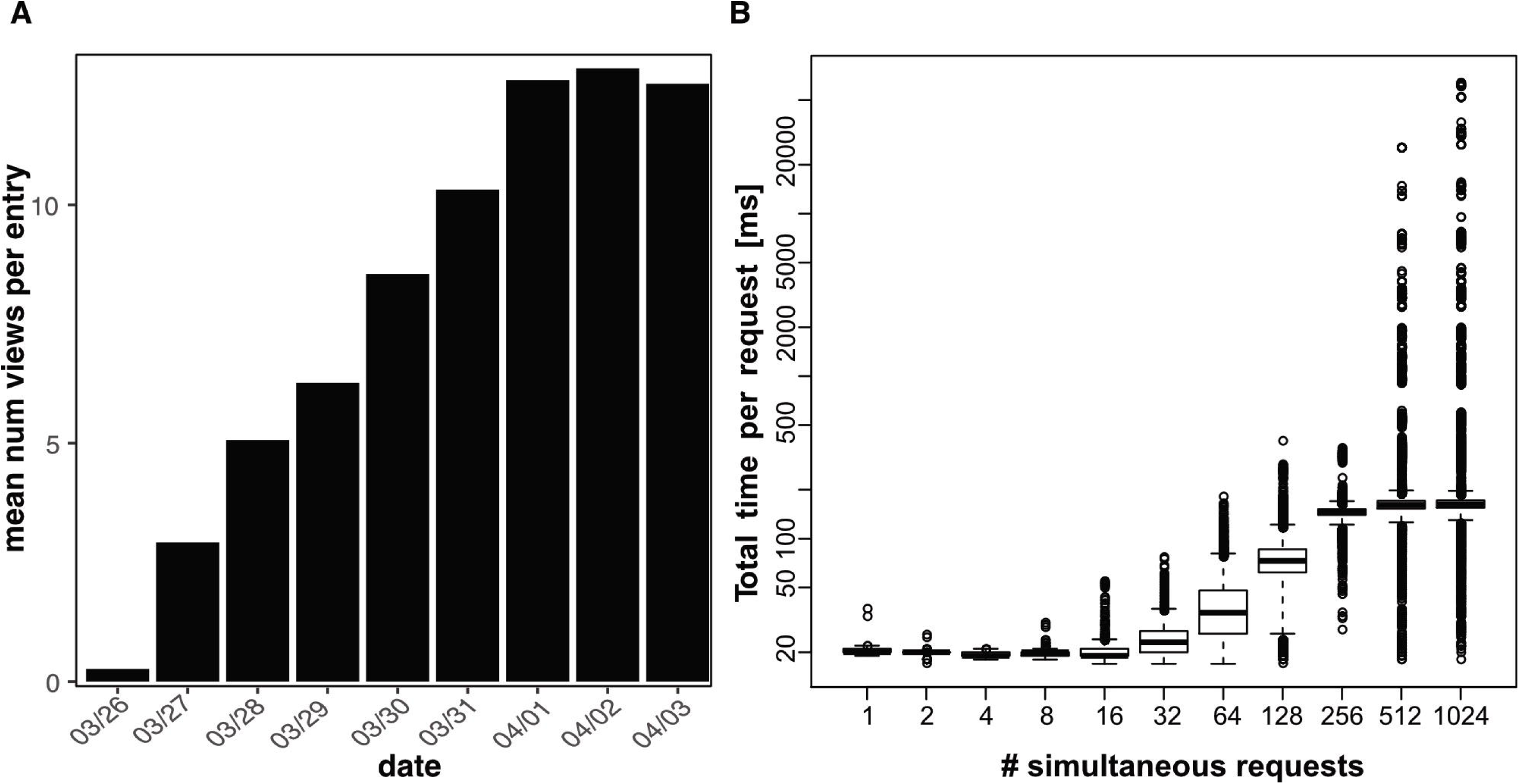
Requests to the CTest server. (A) Since its introduction in the routine of the University Hospital Ulm (Germany), a mean of 12 views per test was recorded (for March 26th only the last 3.5 hours were recorded). (B) The server can respond within 200 ms to >80% of requests when doing up to 1024 simultaneous requests. In this simulation, requests were made using a single machine and a single network connection.

Additionally, we checked its performance and robustness in load tests (Fig 4B and S1-S6 Figs). Our CTest server can respond within 200 ms to over 80% of requests and within 500 ms to over 90% when doing up to 1024 simultaneous requests (Fig 4B). Furthermore, we performed stress tests with available and unavailable tracking numbers (S1-S6 Figs). Based on these tests, we are confident that CTest is well-suited to the demands of rapid testing even if deployed in an ad hoc manner on standard hardware. Its platform independence allows its deployment on a wide variety of (existing) infrastructures.

Another feature of CTest is its functionality for error reporting and statistics. A dedicated “reporter” account is required to access the reporting data. Here, all data is provided in the machine-readable JSON format and the path “/reports/list” can be accessed to get a list of information and error reports. The included information is about the successful backup runs and successful test status imports from CSV files. Also, the number of test results views per day can be accessed at “/reports/views”. General system information such as memory consumption and CPU usage are available at “/reports/system”. Furthermore, the sample collection dates are available “/reports/test-dates” for analysis.

We present CTest with a web interface. In addition to its web interface, CTest facilitates the access of third-party software (apps), if the testee decides to use these. These apps can query the status using the tracking link with appended ?app=true which returns only the test status “negative” or “in progress” instead of the complete HTML document.

## Discussion

The primary goal of this approach was to reduce the burden of clinical staff in the COVID-19 crisis. Furthermore, we wanted to empower the testees to obtain their results in a facile and easy to access way while at the same time ensuring efficient and almost instantaneous and exclusive communication.

Speedy communication is essential in the current crisis, as virus carriers can be infectious before first symptoms arise and as tested people are worried until they know the result of their test [5,6]

Not knowing the outcome might tempt testees not to act accordingly and thus increase the risk of infecting others. Therefore, CTest might help to contribute to reducing virus spreading and also can help to reduce the mental stress of the testees.

First analyses of CTest in the clinical routine showed a mean of 12 queries per performed test. Even half of that number of telephone inquiries would lead to communication overload. Consequently, the introduction of CTest into the clinical routine at the University Hospital Ulm could achieve our primary goal of reducing the burden on the clinical staff. Beneficial for this purpose was the open-source license of TraqBio and its clean and simple setup. This made it possible to implement and integrate the CTest system within a very short period (4 days) into the clinical workflow. Another advantage is that users do not have to create an account to request their test results.

Currently, CTest is specialized for the query of COVID-19 test results and their status. However, it can be adapted with a moderate effort to other queries or the distribution of different types of test results. Even the integration of the CTest system into apps is possible if the html view presented here is not desired. For this purpose, the token “?app=true” has to be added after the tracking number.

Tracking numbers for test results are created based on non-personalized order numbers. Thus, CTest does not use or store personal data that allows identification of people being tested. Based on this implementation, we address the challenges of big data in personalized medicine [7] and respect the German and European data protection law. Clinical or laboratory information systems are often closed-source, and development, adaption and integration of new interfaces can be time-consuming. As exploits allow access to different types of sensitive data, we developed an independent, stand-alone software solution without storing personalized data.

We provide two possibilities (web page and QR-code) to query the status of the test results. The fact that around 30% of users query their test results with a web browser encourages us in this direction. CTest has already been successfully integrated into the clinical workflow at the University Hospital Ulm to keep tested persons updated.

## Data Availability

A software is freely available from https://github.com/sysbio-bioinf/CTest under the Eclipse Public License v2.0 (EPL2).

## Acknowledgements

We thank the COVID-19 task force of the University Hospital Ulm, Udo X. Kaisers, CEO of the University Hospital Ulm, the Department of Clinical Chemistry, the Comprehensive Cancer Center Ulm, the ZIK and all our supporters at University Medicine Ulm (Martin Loose, Thomas Baur, Franz Jobst, Robert Mahnke, and others), who supported the development of the tool and allowed its fast integration into the clinical processes. We also thank Nensi Ikonomi, Yessin Yahia, William Ferdinand, Cagatay Günes, and Xenia Kirgis for their help translating the texts. The rapid implementation, deployment and integration was also made possible by support from different digitization initiatives. KAK, HAK and OK acknowledge, funding from the Germany Federal Ministry of Education and Research (BMBF) as part of the DIFUTURE project (Medical Informatics Initiative, grant numbers 01ZZ1804I and 01ZZ1804D). OK and HAK acknowledge funding from the Ministry of Science and Art Baden-Württemberg (Zentrum für Innovative Versorgung, ZIV). OK and TS acknowledge funding from the Ministry of Social Affairs of the state of Baden-Württemberg (Zentren für Personalisierte Medizin, ZPM), HAK also acknowledges funding from the German Science Foundation (DFG, grant number 217328187).

## Availability

The source code, documentation, and an installation guide is freely available from https://github.com/sysbio-bioinf/CTest under the Eclipse Public License v2.0 (EPL2).

## Author contributions

Funding acquisition: KAK, OK, PK, TS, HAK.

Project administration: HAK.

Software: GV, AF, TG, JMK, AG, JDS.

Supervision: HAK.

Visualization: GV, AF, JDS, SDK.

Writing – original draft: JDS, SDK, AF, PK, OK, GV, HAK. Writing – review & editing: JDS, GV, FK, KAK, SDK, OK, TS, HAK.

## Supporting information

**S1 Fig. Stress test with available URLs**. Queries were measured for 758 available database entries (available_urls) in the CTest system, which were accessed 1516 times sequentially (ordered) and randomly (random). Furthermore, tests were performed for the caching function of the browser (1filePerRequest) and complete reloading (23or1filesPerRequest) of the page.

**S2 Fig. Stress test with non-available URLs**. Queries were measured for 758 non-available database entries (notavailiable_urls) in the CTest system, which were accessed 1516 times once sequentially (ordered) and randomly (random). Furthermore, tests were performed for the caching function of the browser (1filePerRequest) and complete reloading (23or1filesPerRequest) of the page.

**S3 Fig. Stress test with a combination of available and non-available URLs**. Queries were measured for a mixed set of database entries (interleaved_urls) including 758 available CTest entries and the same number non-available database entries which were accessed 1516 times once in sequence (ordered) and randomly (random). Furthermore, tests were performed for the caching function of the browser (1filePerRequest) and complete reloading (23or1filesPerRequest) of the page. This means that once unchanged files are not reloaded (caching), whereas in the other scenario, all required files and displayed images (e.g. flag-graphics or css) are reloaded. For the second scenario, one file is loaded if the tracking number is not available and 23 files if the tracking number is available.

**S4 Fig. Stress test with one available URL**. Queries were measured for one available URL (one_available_url) in the CTest system, which was accessed 1516 times once in sequence (ordered) and randomly (random). Furthermore, tests were performed for the caching function of the browser (1filePerRequest) and complete reloading (23or1filesPerRequest) of the page. This means that once unchanged files are not reloaded (caching), whereas in the other scenario, all required files and displayed images (e.g. flag-graphics or css) are reloaded. For the second scenario, one file is loaded if the tracking number is not available and 23 files if the tracking number is available.

**S5 Fig. Stress test with one non-available URL**. Queries were measured for one not available URL (one_notavailable_url) in the CTest system, which was accessed 1516 times once in sequence (ordered) and randomly (random). Furthermore, tests were performed for the caching function of the browser (1filePerRequest) and complete reloading (23or1filesPerRequest) of the page. This means that once unchanged files are not reloaded (caching), whereas in the other scenario, all required files and displayed images (e.g. flag-graphics or css) are reloaded. For the second scenario, one file is loaded if the tracking number is not available and 23 files if the tracking number is available.

**S6 Fig. Stress test with two URL**. Queries were measured for two URLs (1 available and one not available; two_interleaved_url) in the CTest system, which was accessed 1516 times once in sequence (ordered) and randomly (random). Furthermore, tests were performed for the caching function of the browser (1filePerRequest) and complete reloading (23or1filesPerRequest) of the page. This means that once unchanged files are not reloaded (caching), whereas in the other scenario, all required files and displayed images (e.g. flag-graphics or css) are reloaded. For the second scenario, one file is loaded if the tracking number is not available and 23 files if the tracking number is available

